# Catching a resurgence: Increase in SARS-CoV-2 viral RNA identified in wastewater 48 hours before COVID-19 clinical tests and 96 hours before hospitalizations

**DOI:** 10.1101/2020.11.22.20236554

**Authors:** Patrick M. D’Aoust, Tyson E. Graber, Elisabeth Mercier, Danika Montpetit, Ilya Alexandrov, Nafisa Neault, Aiman Tariq Baig, Janice Mayne, Xu Zhang, Tommy Alain, Mark R. Servos, Nivetha Srikanthan, Malcolm MacKenzie, Daniel Figeys, Douglas Manuel, Peter Jüni, Alex E. MacKenzie, Robert Delatolla

**Author notes:** Corresponding author: **Dr. Robert Delatolla**, Work.

## Abstract

Curtailing the Spring 2020 COVID-19 surge required sweeping and stringent interventions by governments across the world. Wastewater-based COVID-19 epidemiology programs have been initiated in many countries to provide public health agencies with a complementary disease tracking metric and facile surveillance tool. However, their efficacy in prospectively capturing resurgence following a period of low prevalence is unclear. In this study, the SARS-CoV-2 viral signal was measured in primary clarified sludge harvested every two days at the City of Ottawa’s water resource recovery facility during the summer of 2020, when clinical testing recorded daily percent positivity below 1%. In late July, increases of >400% in normalized SARS-CoV-2 RNA signal in wastewater were identified 48 hours prior to reported >300% increases in positive cases that were retrospectively attributed to community-acquired infections. During this resurgence period, SARS-CoV-2 RNA signal in wastewater preceded the reported >160% increase in community hospitalizations by approximately 96 hours. This study supports wastewater-based COVID-19 surveillance of populations in augmenting the efficacy of diagnostic testing, which can suffer from sampling biases or timely reporting as in the case of hospitalization census.

## 1. Introduction

The COVID-19 global pandemic has profound impacts on day-to-day life through sickness, death and also the economic and social effects of lockdowns, closures and curfews put in place by local governments to control and limit the community spread of the disease. COVID-19 surveillance approaches invoked by nations around the world include case ascertainment of individual patients and include nucleic acid-based tests, serological tests and contact tracing. The objective of this study is to investigate early identification of COVID-19 infection resurgence in Ottawa (Ontario, Canada), using SARS-CoV-2 viral RNA signal in primary sludge wastewater.

Wastewater-based epidemiology (WBE) is a potential surveillance tool to obtain a rapid measure of COVID-19 infection in the population at large. Fecal shedding of SARS-CoV-2 viral particles has been shown to occur before, during and after active COVID-19 infection, for periods ranging from a few days to several weeks (Chen et al., 2020; Cheung et al., 2020; Xing et al., 2020; Xu et al., 2020), with current literature suggesting that children may shed viral particles longer than adults (Li et al., 2020; Ma et al., 2020). Fecal shedding of SARS-CoV-2 viral particles in stool occurs in patients infected with COVID-19 and appears to be independent of the presence of SARS-CoV-2 viral particles in the upper respiratory tract. As WBE initiatives rely on the fecally shed viral particles (Collivignarelli et al., 2020; Lodder and de Roda Husman, 2020), an understanding of the predictive ability of this epidemiological metric is necessary for public health units to appropriately action this tool (Hart and Halden, 2020; Hill et al., 2020; Thompson et al., 2020). In particular, studies to date that demonstrate the potential for WBE to identify the onset of community infection prior to clinical testing have largely been applied to identifying first wave of the disease in community (Kaplan et al., 2020; La Rosa et al., 2020; Medema et al., 2020; Nemudryi et al., 2020; Peccia et al., 2020; Sherchan et al., 2020; Vallejo et al., 2020; Wu et al., 2020). Due to limited clinical testing in many countries during the onset of COVID-19 and limited use of wastewater-based SARS-CoV-2 surveillance, it remains unclear whether WBE can have a role in identifying COVID-19 resurgences. The objectives of this study are to establish a relationship between: i) increases in SARS-CoV-2 RNA signal in wastewater, ii) increases in the number of new COVID-19 positive patient cases in the community, and iii) increases in the number of new hospitalizations of COVID-19 positive patients in the community.

## 2. Materials and methods

### 2.1. Characteristics of the City of Ottawa’s water resource recovery facility

Primary clarified sludge samples were harvested from the City of Ottawa’s Robert O. Pickard Environmental Centre, in Ottawa, ON, which services approximately 1.0M residents of the national capital region. The hydraulic residence time of the Ottawa sewershed ranges from 2 hours to 35 hours, with an average residence time of approximately 12 hours. The Ottawa water resource recovery facility (WRRF) has an average daily flow of 435,000 m^3^ per day. The facility is comprised of preliminary treatment consisting of coarse and fine screening and grit chambers for the removal of larger particles. The primary treatment units consist of rectangular primary clarifiers and the secondary treatment is a conventional activated sludge system operating without nitrification. Finally, chlorination is used as a disinfection method followed by dechlorination prior to discharge to receiving waters.

### 2.2. Sample collection, concentration, extraction and quantification

24-hour composite samples of primary clarified sludge (PCS) were collected from the Ottawa WRRF at a frequency of every 2 days for a six-week period, from June 20^th^, 2020 until August 4^th^, 2020. Concentration of samples, extraction of nucleic acids and quantification of SARS-CoV-2 N1 and N2 gene regions were performed according to D’Aoust et al. (2020). It has been previously observed that normalization of SARS-CoV-2 viral copies with pepper mild mottle virus (copies SARS-CoV-2/copies PMMoV) was used to normalize viral SARS-CoV-2 signal for variations in wastewater chemico-physical characteristics, plant flows, wastewater solids organic/inorganic ratio and PCR amplification (D’Aoust et al., 2020). Evaluation of viral recovery efficiency was also performed as per D’Aoust et al. (2020); with the data in this study not being corrected for recovery efficiency. Dilution tests were periodically performed to determine if inhibition was present in the samples. The method’s limit of detection for the N1 and N2 gene regions was determined by determining the concentration at which a detection rate of ≥ 95% (<5% false negatives) was obtained, as per the MIQE recommendations (Bustin et al., 2009). Additionally, samples were discarded if they did not meet the following requirements: 1) standard curves are linear (R^2^ ≥ 0.95), 2) the copies/well are found in the linear range of the standard curve, and 3) the primer efficiency was between 90%-130%. Additionally, all samples were analyzed in triplicate and samples with values greater than 2 standard deviations away from the mean were discarded.

### 2.3. COVID-19 epidemiological data

#### Clinical epidemiological data

During the study period, there were two main methods of COVID-19 surveillance. The first method was to observe the daily number of new confirmed COVID-19 infections, determined via laboratory-confirmed SARS-CoV-2 tests using reverse transcriptase-quantitative polymerase chain reaction (RT-qPCR) that targeted the RNA-Dependent RNA polymerase for multiple genes. Tests were classified as positive for SARS-CoV-2 infection if at least one of any of the two gene “targets” were detected (Government of Ontario, 2020). There was a notable increase in COVID-19 testing in Ottawa during early June with an average daily testing increase from approximately 270 tests per day to approximately 800 tests per day. As a result of increased testing resources, testing was predominantly performed at dedicated community clinics, physician offices and in hospitals, retirement homes and long-term care facilities.

The second method used for COVID-19 surveillance was the daily hospital census counts, defined as the number of patients residing in Ottawa with confirmed COVID-19 infections at midnight to any of Ottawa five acute care hospitals that accepted COVID-19 patients. The average length of stay during the study period was 19.5 days, interquartile range 22 days.

Additional methods used for surveillance of COVID-19 infection is are the number of daily cases and test positivity. These metrics are commonly communicated to the public. COVID-19 test positivity is the proportion of positive COVID-19 cases as a percent of all individuals tested for COVID-19 on a specific date. Confirmed COVID-19 cases and positivity were examined using daily counts based on the date of the test reported for Ottawa residents and seven-day mid-point average.

### 2.4. Statistical analysis

In order to test for significance and for strength of correlation between SARS-CoV-2 RNA signal and epidemiological metrics, a student’s t-test and Pearson’s correlation analyses were performed, with a *p*-value of 0.05 or lower signifying significance.

To evaluate if a lag existed between the appearance of increased SARS-CoV-2 RNA signal in wastewater and epidemiological metrics, time-step analyses were also performed where the correlations between viral RNA signal and epidemiological metrics were offset by a period of 1 to 7 days. The time-step analysis was performed across three discreet time-periods: i) pre-resurgence and resurgence, ending after the resurgence (June 21^st^ to July 21^st^), ii) pre-resurgence and post-resurgence, extending to the end of the data set (June 21^st^ to July 25^th^) and iii) full data period (June 21^st^ to August 4^th^).

## 3. Results, discussion and implications

### 3.1. Trend and peak of wastewater SARS-CoV-2 RNA viral level and other COVID-19 surveillance metrics

All conventional COVID-19 surveillance measures increased during the study period from a low level at the beginning of the study. The number of daily cases and percent positive cases was highest on July 19, with a 326% relative increase from the preceding 7-day period. Meanwhile, current hospitalized patient cases were at their highest on July 31-Aug 2 with a 168% increase compared to the week of July 13 (Figure 2 a-d).

**Figure 1:**
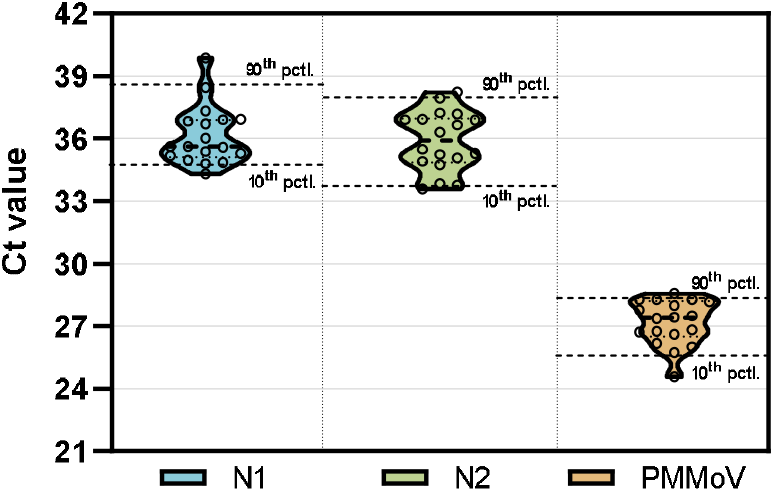
Ct values of the SARS-CoV-2 N1 and N2 gene regions, and the PMMoV normalization gene (1:10 dilution shown), outlining the greater stability of PMMoV, justifying its use as a normalization gene. The 10^th^ and 90^th^ percentile are displayed, along with the median (dotted line inside shapes).

**Figure 2:**
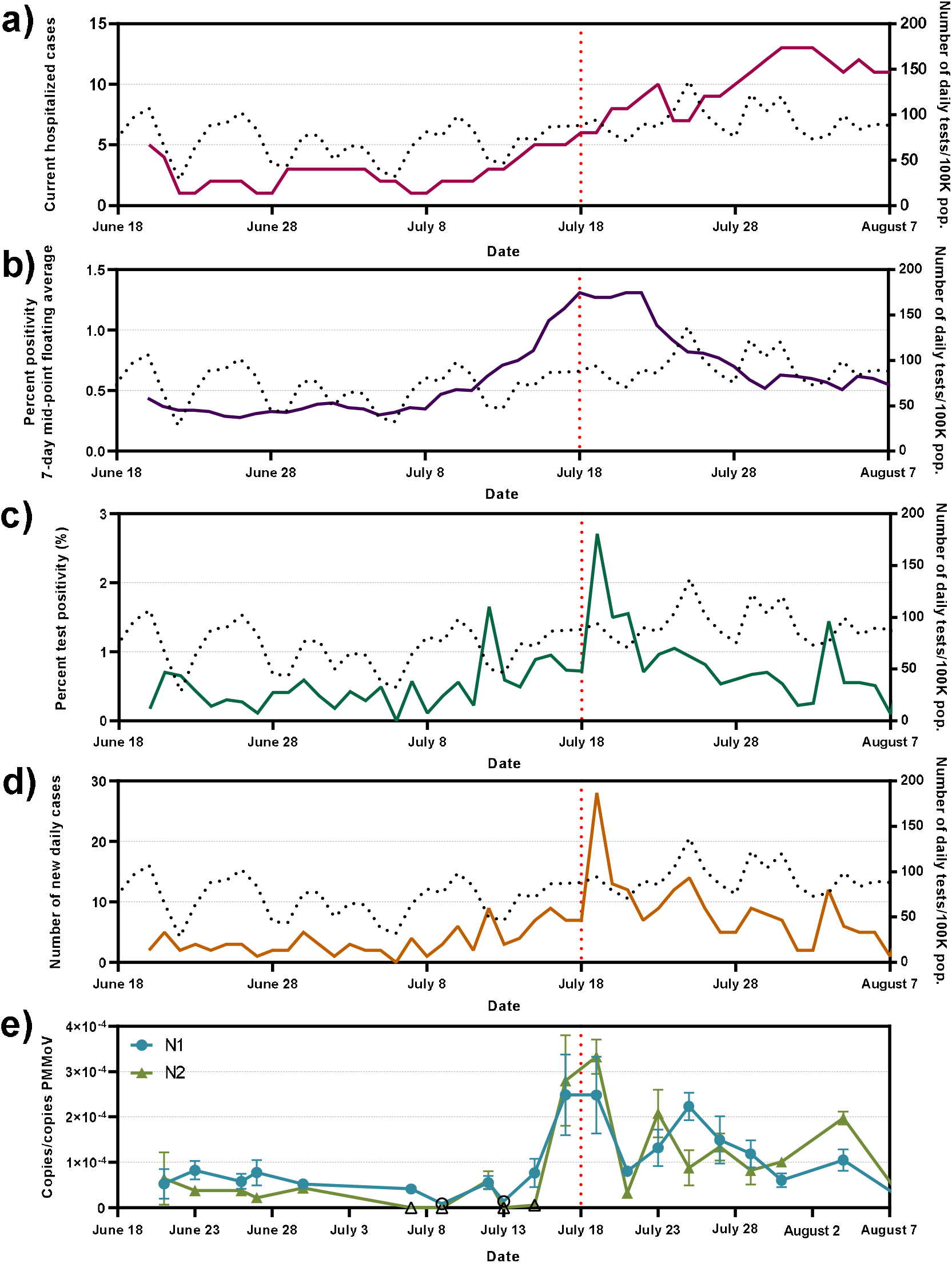
Epidemiological metrics and SARS-CoV-2 viral concentrations over the study period; a) COVID-19-caused hospitalizations, b) percent test positivity (7-day mid-point floating average), c) percent test positivity and d) number of new daily cases, along with e) wastewater SARS-CoV-2 N1 and N2 gene copies/PMMoV gene copies. Open data points signify points which were below the limit of quantification.

PMMoV normalized SARS-CoV-2 viral copies, a metric outlined in an earlier publication (D’Aoust et al., 2020), normalizes SARS-CoV-2 viral copies by the number of copies of PMMoV in wastewater, and is effective due to the great temporal stability of PMMoV, as demonstrated in Figure 1, below. Additionally, dilution tests demonstrated that the sample PCR was not inhibited.

Similarly to other epidemiological metrics, the PMMoV normalized viral signal in Ottawa’s wastewater was also shown to be low at the beginning of the study period through to the week of July 13 with both N1 and N2 regions below 1.00×10^−4^ copies/copies PMMoV. There was a 450% relative increase (*p*<0.05) in N1 and 440% increase (p<0.05) in N2 viral regions on July 17 to 19 compared to July 13 to 15. The absolute increase of 2.03×10^−4^ and 3.01×10^−4^ copies/copies PMMoV was reported during this period of increase for N1 and N2 gene regions, respectively (Figure 2e). The wastewater-based epidemiological metric in this study was not corrected for recovery percentages.

### 3.2. Correlation between surveillance metrics

Positive correlations were observed (Figure 2 c, d and e) between the normalized viral RNA signal and both the number of new daily positive COVID-19 cases (N1: R = 0.673, *p*<0.001; N2: R = 0.648, *p*<0.001) and clinical testing percent positivity (N1: R = 0.468, *p*<0.001; N2: R = 0.404, *p*<0.001). These findings are in agreement with reported observations from previous studies (D’Aoust et al., 2020; Nemudryi et al., 2020; Peccia et al., 2020). This study, as opposed to an earlier study in the same region (D’Aoust et al., 2020), demonstrates that strong correlations do in fact exist between viral RNA signal and the epidemiological metric of clinical daily new COVID-19 cases, as supported by several other studies (Hart and Halden, 2020; Michael-Kordatou et al., 2020; Trottier et al., 2020; Wu et al., 2020). The moderate correlations between viral RNA signal and clinical daily new COVID-19 cases in previous work was largely attributed to inadequate resources to achieve a developed clinical testing framework early in the pandemic (lower daily # of tests). In this study, a strong testing regiment was deployed in Ottawa and hence produced a more reliable daily new COVID-19 cases metric across this study period. A moderately weak positive correlation exists between N1 and N2 SARS-CoV-2 PMMoV-normalized RNA signal and COVID-19-caused hospitalizations (R = 0.347, *p*<0.001 and R = 0.464, *p*<0.001 for N1 and N2, respectively; Figure 2 a and e).

### 3.3. Temporal association between surveillance metrics

A visual comparison of clinical testing percent positivity and the number of new daily positive COVID-19 cases compared to the PMMoV-normalized RNA signal shows that the viral RNA signal predates both the clinical testing percent positivity and new daily positive COVID-19 cases epidemiological data. A times-step correlation shows the strongest correlation for a two-day step between SARS-CoV-2 viral RNA signal and both the number of new daily positive COVID-19 cases and the clinical testing percent positivity (N1: R = 0.703, p<0.001; N2: R = 0.721, p<0.001N1: R = 0.703, p<0.001; N2: R = 0.714, p<0.001) (Table 1). The strong correlation for hospitalized cases is observed after a time-step of four days (N1: R = 0.741, p<0.001; N2: R = 0.767, p<0.001). The precedence of viral RNA signal vs. other epidemiological metrics has been reported recently by other earlier studies (Chavarria-Miró et al., 2020; Kumar et al., 2020; Peccia et al., 2020).

**Table 1:** Time-step analyses of correlations (Pearson’s R) between normalized SARS-CoV-2 viral RNA signal (copies/copies PMMoV) and 7-day rolling average percent positivity, test percent positivity and daily new cases epidemiological metrics.

|  | Daily offset | Hospitalized cases |  | 7-day rolling average test percent positivity |  | Test percent positivity |  | Daily new cases |  |
| --- | --- | --- | --- | --- | --- | --- | --- | --- | --- |
|  |  | N1 | N2 | N1 | N2 | N1 | N2 | N1 | N2 |
| June 21 - July 21 | 0 | 0.488 | 0.502 | 0.675 | 0.662 | 0.551 | 0.635 | 0.680 | 0.754 |
|  | 1 | 0.599 | 0.578 | 0.662 | 0.647 | 0.634 | 0.723 | 0.642 | 0.692 |
|  | 2 | 0.532 | 0.487 | 0.666 | 0.679 | 0.853 | 0.821 | 0.810 | 0.765 |
|  | 3 | 0.747 | 0.754 | 0.556 | 0.593 | 0.191 | 0.208 | 0.354 | 0.310 |
|  | 4 | 0.741 | 0.767 | 0.415 | 0.457 | 0.338 | 0.332 | 0.265 | 0.237 |
|  | 5 | 0.512 | 0.487 | 0.301 | 0.341 | 0.144 | 0.140 | 0.375 | 0.405 |
|  | 6 | 0.534 | 0.490 | 0.104 | 0.167 | -0.031 | 0.023 | 0.107 | 0.190 |
|  | 7 | 0.375 | 0.361 | -0.012 | 0.077 | -0.040 | 0.070 | 0.078 | 0.178 |
| June 21 - July 25 | 0 | 0.539 | 0.551 | 0.659 | 0.688 | 0.505 | 0.612 | 0.707 | 0.712 |
|  | 1 | 0.689 | 0.562 | 0.631 | 0.653 | 0.637 | 0.747 | 0.662 | 0.709 |
|  | 2 | 0.634 | 0.489 | 0.623 | 0.667 | 0.724 | 0.794 | 0.699 | 0.783 |
|  | 3 | 0.786 | 0.695 | 0.488 | 0.576 | 0.128 | 0.203 | 0.277 | 0.346 |
|  | 4 | 0.790 | 0.690 | 0.324 | 0.437 | 0.264 | 0.277 | 0.245 | 0.192 |
|  | 5 | 0.632 | 0.483 | 0.188 | 0.303 | 0.114 | 0.108 | 0.378 | 0.354 |
|  | 6 | 0.645 | 0.482 | 0.038 | 0.118 | -0.073 | 0.008 | 0.102 | 0.202 |
|  | 7 | 0.531 | 0.408 | -0.074 | 0.015 | -0.157 | 0.041 | -0.032 | 0.163 |
| Whole data set | 0 | 0.347 | 0.470 | 0.644 | 0.626 | 0.469 | 0.527 | 0.673 | 0.648 |
|  | 1 | 0.453 | 0.484 | 0.606 | 0.602 | 0.634 | 0.668 | 0.654 | 0.634 |
|  | 2 | 0.429 | 0.424 | 0.593 | 0.603 | 0.703 | 0.714 | 0.703 | 0.721 |
|  | 3 | 0.556 | 0.565 | 0.448 | 0.491 | 0.027 | 0.081 | 0.182 | 0.375 |
|  | 4 | 0.603 | 0.586 | 0.307 | 0.360 | 0.233 | 0.278 | 0.223 | 0.158 |
|  | 5 | 0.497 | 0.477 | 0.165 | 0.234 | 0.079 | -0.034 | 0.306 | 0.206 |
|  | 6 | 0.546 | 0.500 | 0.020 | 0.064 | -0.091 | 0.021 | 0.074 | 0.195 |
|  | 7 | 0.468 | 0.443 | -0.080 | -0.119 | -0.074 | 0.005 | -0.025 | 0.178 |

This observation is supported by, and confirms earlier studies (Chavarria-Miró et al., 2020; Kumar et al., 2020; Peccia et al., 2020). Recent studies proposed that viral SARS-CoV-2 RNA data may in fact precede hospital admissions by 1 to 4 days in primary sludge (Figure 3) (Peccia et al., 2020), and 6-14 days in raw wastewater (Kumar et al., 2020). This could signify that hospitalizations may begin peaking four days after an increased viral RNA signal, with the caveat that hospitalizations rates may be significantly higher when viral RNA signal is the result from shedding of an at-risk population group (young infants, elderly, immunocompromised, etc.). In addition, the RNA signal post-resurgence indicates that the resurgence events in viral RNA signal serves to weaken the correlation of subsequent peaks with clinical testing metrics. It is hypothesized that early and later infections and overlapping shedding events of duration and intensity convolute peaks within a period of time following the initial new peaks/surges in COVID-19 infections. Nonetheless, the RNA epidemiological metric of copies/copies PMMoV may become extremely valuable to public health units in their preparation for the next COVID-19 wave.

**Figure 3:**
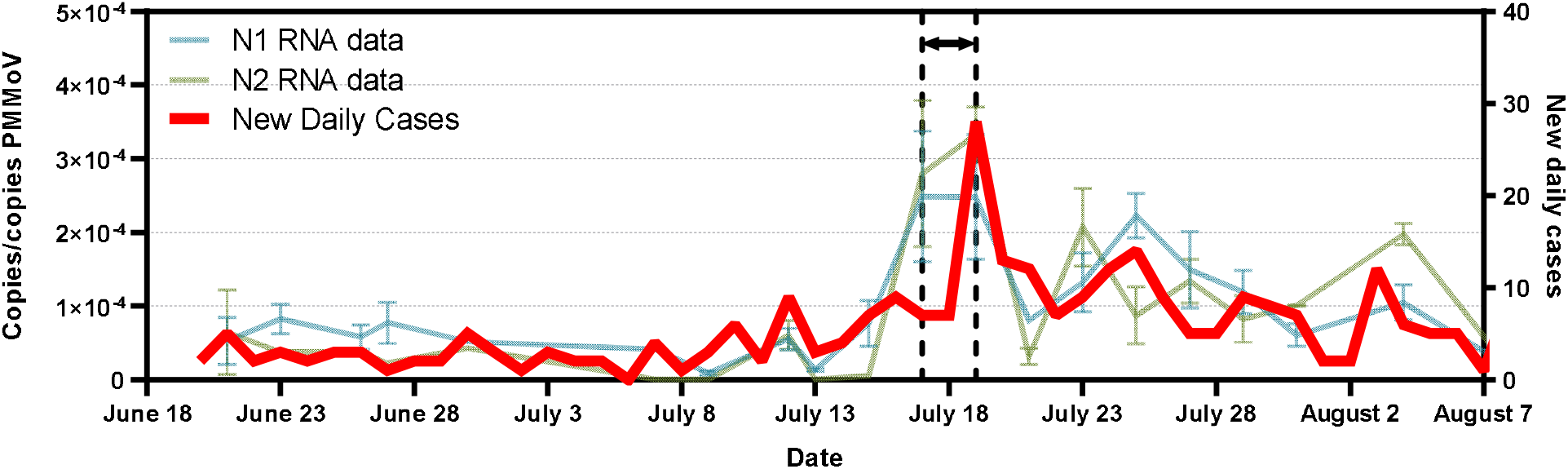
Figure of the new daily cases epidemiological metric (in red) with SARS-CoV-2 viral RNA signal in Ottawa’s primary clarified sludge samples, with the precedence of SARS-COV-2 viral signal in wastewater clearly outlined.

## 4. Conclusions

This study demonstrates that due to the general stability of SARS-CoV-2 RNA measurements in PCS, this fraction of wastewater can reliably be utilized to predict rapid increases or resurgences in COVID-19 cases in the community at large (Alpaslan-Kocamemi et al., 2020; D’Aoust et al., 2020; Peccia et al., 2020), and the results strongly suggests that PCS can be utilized as part of sentinel-type wastewater based epidemiology endeavors at WRRFs.

Advantages of this approach include: i) the analyses in water resource recovery facilities are high-enrollment surveys, which capture a majority of the urban population, regardless of individual’s reluctance to get tested clinically; ii) tests are anonymous and do not pose ethical challenges as opposed to mass clinical testing or targeted sewershed monitoring efforts; iii) data can be used as a confirmation to clinical tests, increases the effectiveness of local public health units and data remains independent of official testing strategies or media sentiment, and iv) provides an exit strategy/path forward for public health units as something to eventually transition to once immunization is available, to cut down on clinical testing costs, while maintaining broad surveillance capabilities. However, limitations to this WBE approach includes the following: i) data obtained only gives high-altitude view of the situation in the community, and it is currently not possible to correlate this data to an actual number of cases in the community; ii) testing protocols must be optimized and retain sensitivity at very-low levels of disease incidence to remain relevant; iii) widespread adoption of WBE requires scale-up of public health units’ current capabilities and/or partnership with private laboratories or research institutions, and iv) increases in resolution (more localized, upstream of water resource recovery facilities, for example) of tests in watersheds may lead to ethical questions for public health unit if it wishes to act upon the data, due to the potential risk for identifying or singling out small subgroups of a population. It is noted however that these issues are not dissimilar to other ethical issues currently existing with other COVID-19 public health endeavors, such as COVID-19 potential exposure notification smartphone applications.

## Data Availability

Data available upon request, and most up to date data available live at www.613covid.ca/wastewater

## Acknowledgements

The authors wish to acknowledge the help and assistance of the University of Ottawa, the Ottawa Hospital, the Children’s Hospital of Eastern Ontario, the Children’s Hospital of Eastern Ontario’s Research Institute, the City of Ottawa, Ottawa Public Health, Public Health Ontario and all their employees involved in the project during this study. Their time, facilities, resources and thoughts provided throughout the study helped the authors greatly. The authors also wish to specifically outline the assistance of Dr. Monir Taha at Ottawa Public Health.

## 6. Supplemental Material

**Supplementary Table 1:**
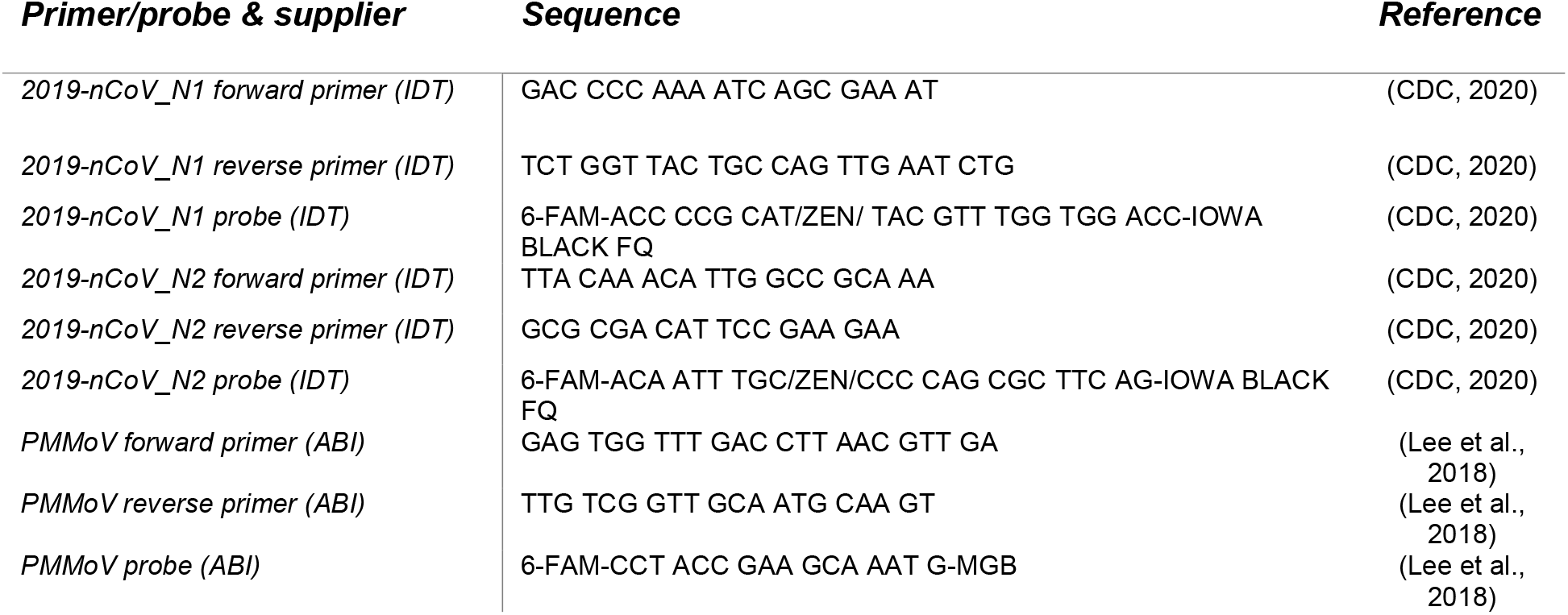
List of PCR primer and probe sets utilized in this study.

